# Design and model choices shape inference of age-varying genetic effects on complex traits

**DOI:** 10.1101/2025.07.01.25330633

**Authors:** Tabea Schoeler, Simon Wiegrebe, Thomas W. Winkler, Zoltán Kutalik

## Abstract

Understanding how genetic influences on complex traits change with age is a fundamental question in genetic epidemiology. Both cross-sectional (between-subject) and longitudinal (within-subject) approaches can contribute to answering this question, but come with distinct strengths and limitations.

Using data from 31 health-related phenotypes in the UK Biobank, we applied a two-stage genome-wide approach to identify genetic variants exhibiting age-dependent effects. To assess the robustness of these findings, we tested for variant-specific results across multiple analytical models, and linked these to differences in key methodological assumptions. Within this framework, we systematically compared the results from cross-sectional gene-by-age interaction models (up to 406,226 individuals) with those from genetic association tests on longitudinal change (up to 83,579 individuals with repeat measurements), and investigated potential sources of bias underlying any observed discrepancies.

We found high concordance in the direction of age-varying genetic effects across the two designs (85.96% of the 57 identified variants), but only moderate agreement in magnitude of effect sizes (Pearson *r* = 0.74). Gene-by-birth year effects, which bias cross-sectional estimates, accounted for the largest proportion of variance in effect size differences across SNPs with age-varying effects between designs (53.1%). Participation bias accounted for an additional 13.3%, while unmodeled non-linear age trajectories contributed minimally to these differences (2.1%).

Overall, our results demonstrate that both cross-sectional and longitudinal designs can yield different estimates of age-varying genetic effects, principally due to cohort confounding and participation bias. As neither approach is immune to bias, we recommend integrating both designs for robust inference, to help minimize bias and more accurately characterize how genetic effects on complex traits change over the lifespan.

## Introduction

A central question in life course and developmental genetic epidemiology is whether—and how—the impact of genetic variation on complex traits changes with age. Understanding age-varying genetic effects provides critical insights into developmental processes, differential aging trajectories, and the aetiology of age-related diseases. Explicit modeling of genome-wide age-varying effects can thus reveal important patterns of diminishing (or intensifying) genetic influence across the life course−patterns that would be missed by standard genome-wide association tests screening for average effects across time.

Detecting and characterizing age-varying genetic effects is not only crucial for elucidating underlying biological mechanisms, but also for ensuring robust inference in genetic epidemiology. For example, age-dependent effects may obscure SNP heritability estimates, contributing to the so-called ‘missing heritability’^1–3^. Furthermore, such varying effects create challenges for an array of methods assuming stable genetic effects across different age groups, including Mendelian randomization^4^, fixed-effects genome-wide meta-analyses^5,6^, or polygenic prediction^7^. Given the documented evidence of varying genetic effects across the lifespan^8,9^, such presence can therefore complicate the interpretation of common genomic outputs.

Traditionally, investigations into age-dependent genetic effects have predominantly utilized cross-sectional gene-by-age interaction tests, indirectly approximating age-related changes by comparing genetic associations across individuals of differing ages^10–14^ (**Figure 1**, A1). In contrast, longitudinal assessments provide a more direct measure of the aging process, by modeling within-individual change over time and its genetic determinants (**Figure 1**, A2). Although longitudinal genome-wide analyses were previously constrained by the lack of repeated measures in large cohorts, the emergence of large-scale prospective biobanks such as the UK Biobank (UKB) now enables the estimation of genetic effects on change at genome-wide scale^15–18^.

**Figure 1.**
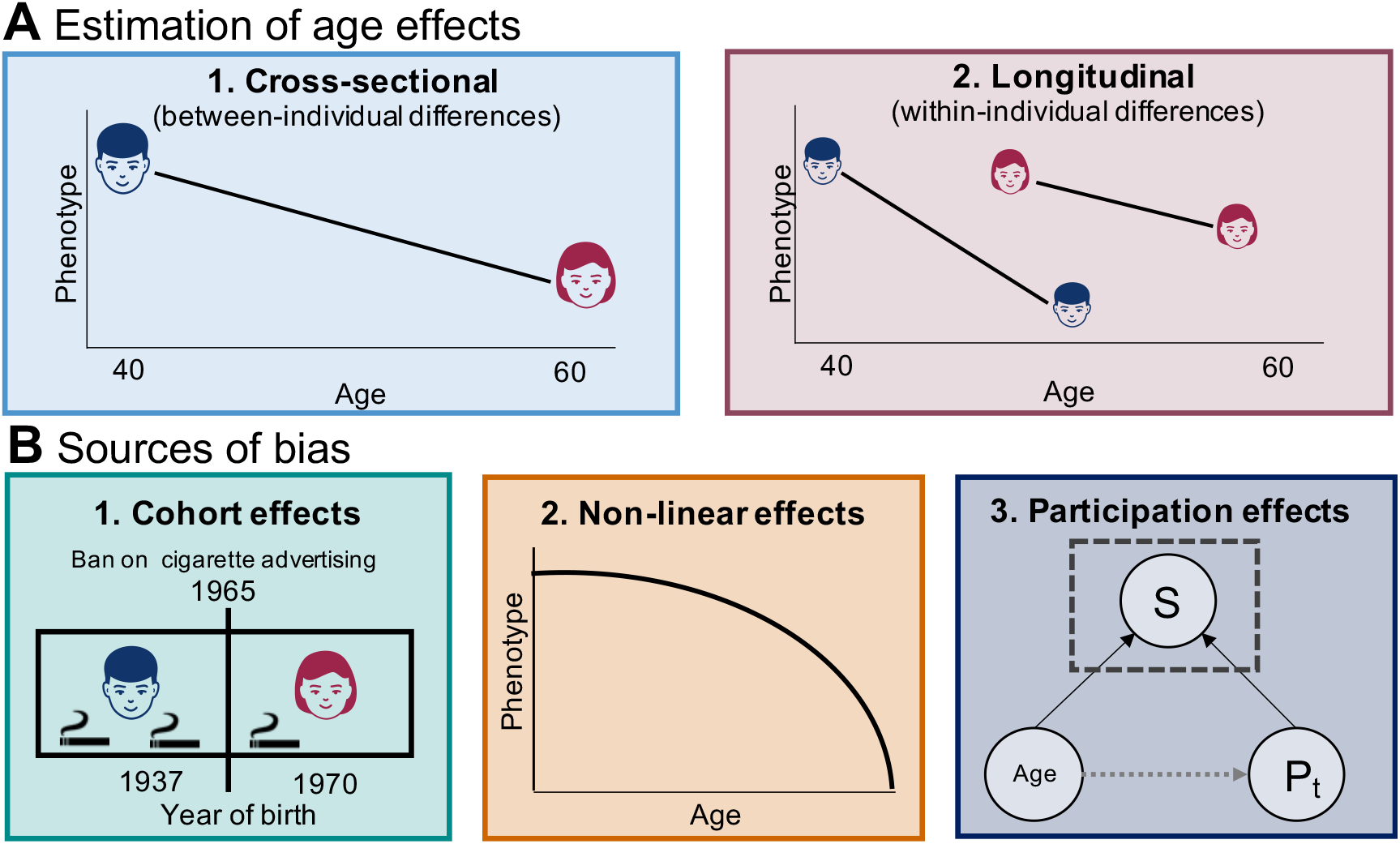
Study framework. **Panel A** illustrates the cross-sectional (between-individual, in blue) and longitudinal (within-individual, in red) design used for the identification and estimation of age and age-varying genetic effects. **Panel B** illustrates possible sources of bias when estimating age and age-varying genetic effects, including (1) birth cohort effects (differences arising from historical or societal changes affecting people born at different times, for example the 1965 UK ban on cigarette advertising reducing cigarette use in younger cohorts), (2) non-linear age effects (where age effects depend on the age of the individual) and (3) selective participation (where selection into the sample, S, depends on age and the phenotype, P, assessed at time point t.)

While both cross-sectional and longitudinal approaches aim to uncover the same underlying biological processes, discrepant findings may arise due differences in model assumptions and sampling features that characterize the two designs. Cross-sectional designs often benefit from significantly larger samples and wider age ranges (e.g., 40–69 years in ~500,000 UKB participants), thereby maximizing statistical power and reducing false negatives^19^. Additionally, they are potentially less susceptible to selective participation (**Figure 1**, B3) than longitudinal designs, in which (age-related) participation bias may be amplified due to selective dropout or survival. Conversely, longitudinal designs, despite smaller sample sizes and shorter follow-up windows (e.g., ~10 years in up to 100,000 UKB participants), provide more robust control against time-invariant sources of confounding, particularly gene-by-birth year (i.e., cohort) interactions (**Figure 1**, B1). For example, cohort-varying genetic effects have been documented in response to societal changes, such as the collapse of the Soviet Union^20,21^ or educational policy reforms^22^−effects that would falsely be interpreted as age-varying genetic effects if tested cross-sectionally. Additionally, longitudinal data with multiple repeated measures enable the detection of non-linear age-related changes, offering deeper insights into age-varying genetic effects that deviate from linear trends (**Figure 1, B2**).

Given the distinct limitations and strengths characterizing cross-sectional and longitudinal designs, it is critical to understand their relative impact on genome-wide findings and inference. In this work, we therefore systematically evaluate age-varying genetic effects estimated from both approaches, identify sources of discrepancy, and provide guidance on their optimal application in genome-wide analyses on complex traits.

## Methods

We included 498,845 UK Biobank individuals aged between 40 and 69 years who were first seen between 2006 and 2010 in one of the 22 assessment centres, corresponding to cohort of individuals born between 1937 and 1970. From the baseline sample, we selected up to 99,459 individuals who were followed-up in repeat assessment(s) carried out between 2012 and 2024 (mean follow-up duration=11 years, ranging from 2 to 18 years). The UKB was approved by the National Health Service North-West Center Research Ethics Committee (REC No. 16/NW/0274) and this research was conducted under application number 16389.

We selected 31 non-binary phenotypes with at least 50, 000 non-missing repeat-measurements per phenotype, tapping into dimensions of cognition (e.g., reaction time), health (e.g., systolic blood pressure, forced expiratory volume), anthropometrics (e.g., BMI, whole body fat-free mass) or lifestyles (e.g., smoking, alcohol consumption). Excluded were all phenotypes with only one single repeat assessment available, as parts of our modelling approach relied on the use of a subset of UKB participants with at least three-time point data to explore possible non-linear age effects (c.f., below). As a result, we included only phenotypes that were reassessed as part of the UKB neuroimaging follow-up assessments (e.g., excluding biomarker measures such as LDL)^23^. Each phenotype *P*_t_ assessed at a given time point *t* was scaled prior to the inclusion in the analytical models 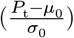, based on their baseline mean (*μ*_0_) and standard deviation (*σ*_0_).

### Cross-sectional and longitudinal age effects

We first performed a series of analyses to assess factors influencing the degree of convergence between age effects obtained from cross-sectional (between-subject) comparisons (*β*_CS_ in A1, c.f., below) and longitudinal (within-subject) comparisons (*β*_L_ in A2), focusing on cohort effects (*β*_C_ in A3), non-linear (quadratic) age effects (*β*_Q_ in A4) and selective participation (weighted *β*_W_ in A5):

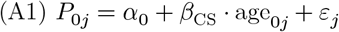

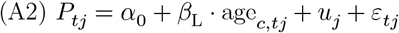

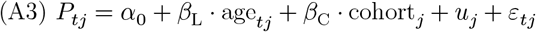

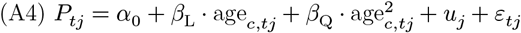

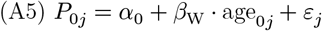

*P*_*tj*_represents a given phenotype at time point *t* (with *t* = 0 representing the phenotype assessed at baseline), for individual *j*. Cross-sectional age effects were obtained from a standard linear regression model (A1), regressing the baseline phenotype (*P*_0*j*_) on baseline age (age_0*j*_), where *α*_0_ denotes the intercept. This model was applied to all UKB participants who had non-missing phenotype values recorded at their initial assessment centre visit, conducted between 2006 and 2010 (up to *N* = 498,845). Models using repeat measurements (A2-A4) were tested by including age as a fixed effect in linear mixed-effects models implemented in *lme4*^24^, where *u*_*j*_denotes the random intercept to account for within-individual clustering and *ε*_*tj*_denotes the residual error. In model (A2), we included only individuals with at least one follow-up assessment (up to *N* = 99,459), and model (A4) was restricted to individuals with at least three assessment waves (up to *N* = 19,374), allowing for the estimation of quadratic age effects. In those two models, age_*c,tj*_represents the person-mean centered age at time *t* for individual 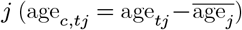, which distinguishes within-individual age effects from between-individual variation25–^28^. Specifically, age centering eliminates bias in the age-estimate that results from systematic variation in mean age across birth years, thereby removing confounding due to cohort effects^26,29,30^. Cohort effects (A3, applied in up to *N* = 99,459 individuals with at least one follow-up assessment) reflect between-individual differences in birth year and were modeled by including both (uncentered) age and year of birth of individual *j* as fixed effects. Age was not centered in this model, as person-mean centering removes between-person variance, thereby precluding the estimation of birth year effects^26,29^. To assess non-linear age effects over time (A4), analyses were restricted to individuals with at least three assessment waves, allowing for the estimation of quadratic age effects. Finally, the impact of selective participation was evaluated by comparing age effects across samples with varying levels of representativeness: Inverse probability weighting (A5) implemented in the *survey*^31^package was applied to obtain weighted age coefficients (*β*_W_) adjusted for selective baseline participation, using previously derived sampling weights^32^. These weighted effects were then compared to those from the unweighted baseline and prospective sample, to assess whether age effects vary as a function of sample representativeness.

To assess the relative contribution of cohort, non-linear, and participation effects to discrepancies between longitudinal (*β*_L_) and cross-sectional (*β*_CS_) age estimates, we modeled the effect size differences as a function of these factors (*β*_L_− *β*_CS_~ *α*_0_ + *α*_1_ ⋅ *β*_C_+ *α*_2_ ⋅ *β*_Q_+ *α*_3_ ⋅ *β*_P_). Here, *β*_P_ indexes participation effects, defined as the difference in cross-sectional age estimates obtained from a representative (weighted estimate, *β*_W_ in model A5) versus a less representative (unweighted estimate, *β*_C_ in A1, obtained in the prospective sample) (*β*_P_= *β*_W_− *β*_C_). The proportion of variance explained by each predictor (*R*^2^) was calculated using the ‘lmg’ method in the *relaimpo*^33^package, following the approach developed by Lindeman, Merenda, and Gold (1980)^34^.

Because the estimates for each factor (*β*_C_, *β*_Q_, *β*_P_) were derived from different subsets of UKB data with varying sample sizes, some are subject to larger estimation error, potentially leading to an underestimation of variance components due to regression dilution^35^. To account for this bias, we calculated the dilution ratio (*λ* ∈ [0, 1]) for each factor, where lower *λ* values indicate greater attenuation. Corrected variance contributions 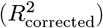 were obtained by dividing the observed *R*^2^-values by *λ* 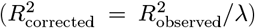. The dilution ratio was computed as 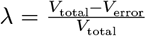, with 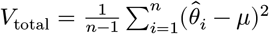 and 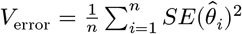. 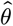 denotes the vector of estimated coefficients and 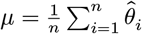 (c.f., **Supplementary Note** for details).

### Identification of age-varying genetic effects

To identify age-varying genetic effects, we implemented a two-step genome-wide strategy that integrates marginal, cross-sectional, and longitudinal models to detect age-varying genetic effects^19^, as illustrated in **Figure 3A**. In the first step, we conducted genome-wide scans for marginal genetic effects (*γ*_*M*_ in model 1), cross-sectional gene-by-age interaction effects (*δ*_CS_ in model 2) and genetic effects on change (*δ*_L_ in model 3).

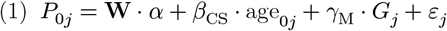

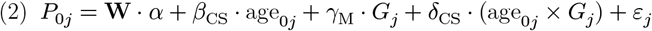

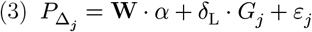

Here, **W** denotes the matrix of covariates (including intercept, the first 20 genetic principal components, genotyping array, and sex), and *α* represents their corresponding coefficients. The outcome for model 3, 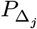, reflects an individual’s annual rate of change in a given phenotype, calculated as the difference between the most recent follow-up and baseline phenotype (*P*_1_ − *P*_0_), divided by follow-up duration 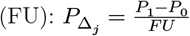. To increase robustness, participants with less than three years of follow-up were excluded from the longitudinal model.

Genome-wide association analyses were conducted using REGENIE (v3.2.6)^36^. First, a wholegenome regression model was fitted using ridge regression to account for local linkage disequilibrium (LD) patterns and relatedness among individuals. Quality-controlled SNPs were included in this step, with filtering criteria of minor allele frequency (MAF) ≥ 1%, Hardy-Weinberg equilibrium *P*-value ≥ 1 × 10^−15^, genotyping rate ≥ 99%, exclusion of SNPs involved in inter-chromosomal LD, and additional LD pruning (*R*^2^ threshold of 0.9, window size of 1,000 markers, step size of 100 markers)^36^. Second, association testing was performed using linear regression models that condition on predictions generated during the first step. This phase utilized the imputed UKB genotype data (version 3, imputed with the Haplotype Reference Consortium panel), considering variants with MAF > 1%. Analyses were restricted to participants of European genetic ancestry, following exclusion of individuals displaying high levels of missingness or extreme autosomal heterozygosity (as defined by UKB^37^). In REGENIE, we used the parameter ‘–interaction age –no-condtl’ to obtain the gene-by-age interaction (*δ*_CS_ in model 2) in addition to the (unconditioned) marginal genetic effects (*γ*_M_ in model 1) within the same iteration. Genome-wide tests on age-related annual change (*δ*_L_ in model 3) were obtained from the standard REGENIE analysis without the specification of the interaction parameter.

In the final step, we identified candidate genetic variants by selecting all LD-independent SNPs that reached genome-wide significance (*P* < 5 × 10^−8^) in any of the three models of marginal effects (*γ*_M_ in model 1), cross-sectional interaction effects (*δ*_CS_ in model 2) and/or effects on change (*δ*_L_ in model 3), following stringent clumping in plink^38^ (–clump-kb 10000, –clump-r2 0.001)^39^. From this set, we selected all Bonferroni-corrected significant variants (discovery threshold: *P* < 0.05/*k*_Eff_) identified in the two models testing for age-varying genetic effects (*δ*_CS_and *δ*_L_, models 2 and 3), where *k*_Eff_ denotes the number of genome-wide variants per trait identified in step 1 (**Figure 3A**). Of note, while the Bonferroni correction using *k*_Eff_ is based on dependent tests (as *k*_Eff_ is derived from candidate variants identified across all three genome-wide models), its application to the age-varying models (cross-sectional and longitudinal) imposes a stricter discovery threshold, thus reflecting a conservative approach. The resulting list of genetic variants exhibiting age-varying effects was then used in downstream analyses evaluating the robustness of these findings.

### Cross-sectional and longitudinal age-varying genetic effects

For the set of identified variants, we first evaluated whether the age-varying genetic effects obtained from cross-sectional and longitudinal analyses were consistent with respect to the direction and interpretation of the effects. To do this, we classified the interaction patterns as attenuation, intensification, or cross-over, based on the relationship between the marginal genetic effect and the age-varying effect. Specifically, attenuation was inferred if marginal and interaction effects had opposite signs (e.g., a positive interaction effect with a negative main effect), whereas intensification was indicated by concordant signs (e.g., both main and interaction effects negative). Effects were labeled as “cross-over” when a significant interaction effect was present in the absence of a significant main effect.

To investigate potential sources of discrepancy between cross-sectional and longitudinal estimates of age-varying genetic effects, we extended the previously described phenotypic models (A1–A5) by incorporating the genetic term (*G*_*j*_), and estimated the relevant parameters for all candidate genetic variants with indicated age-varying effects. Specifically, we compared the age-varying genetic effects obtained from the cross-sectional (*δ*_CS_, model B1) and longitudinal (*δ*_L_, model B2) approach, and fitted a series of additional models designed to assess the influence of variations in genetic effects at the level of cohort differences (*δ*_C_, B3), non-linear (quadratic) age effects (*δ*_Q_, B4), and selective participation (*δ*_W_, B5). The parameters of interest (highlighted in red) were estimated as follows:

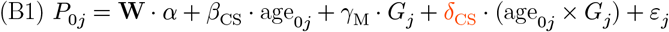

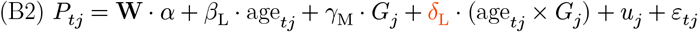

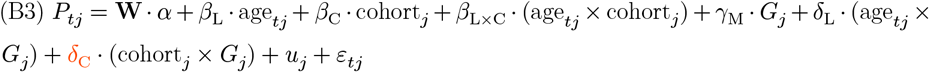

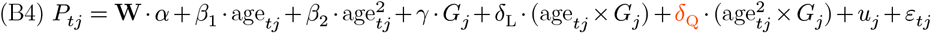

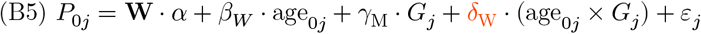

All model parameters were estimated in R in a sample of unrelated UKB participants, applying ordinary least squares regression (B1), linear mixed-effects models (B2–B4), and weighted least squares regression (B5). Of note, the age-by-cohort interaction term in model B3 (*β*_L×C_) was added to appropriately control for age when estimating cohort-varying genetic effects^40^. As in the phenotypic analyses, we quantified the relative contribution of each factor to the observed differences in age-varying genetic effects between the two designs by modeling (*δ*_L_− *δ*_CS_) ~ *α*_0_ + *α*_1_ ⋅ *δ*_C_+ *α*_2_ ⋅ *δ*_Q_+ *α*_3_ ⋅ *δ*_P_, where *δ*_P_ indexes participation effects.

## Results

### Cross-sectional versus longitudinal age effects

To evaluate the convergence between cross-sectional and longitudinal age effects, we focused our analysis on 31 non-binary health-related phenotypes in the UK Biobank, spanning domains relevant to cognition, physiological traits, clinical indicators, anthropometric measures, and lifestyle and behavioural factors. **Figure 2** presents the age-effects obtained from the cross-sectional (A1, up to *N* = 498,845 individuals) and the longitudinal model (A2, up to *N* = 99,459 with at least one follow-up assessment). A summary of all model estimates and corresponding samples sizes is available in **Supplementary Data 1**. Overall, cross-sectional age effects explained 70.57% of the variance in longitudinal estimates. The largest difference in age effects was present for medications (number), where the cross-sectional model substantially overestimated the within-person age effect, with *β*_CS_= 0.03 (standard error, SE = 0.0002) and *β*_L_= 0.01 (SE = 0.0002), due to confounding via cohort effects (further discussed below). Inconsistencies in direction of effects (i.e., opposite sign) were detected for 7 of the 31 included traits, particularly among lifestyle-related traits (e.g., alcohol use, smoking, coffee intake, physical activity).

**Figure 2.**
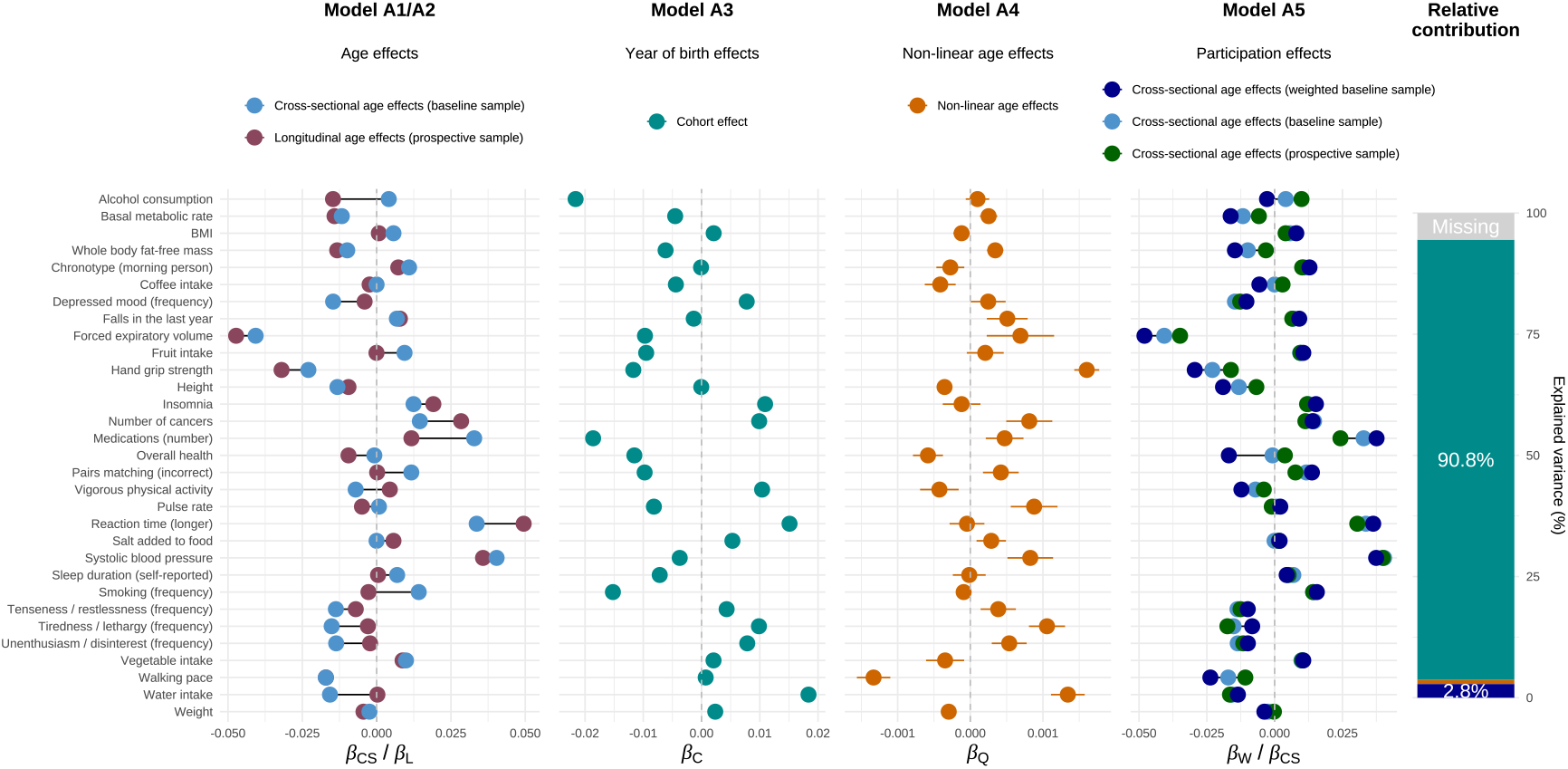
Cross-sectional and longitudinal age effects. **Panel A1/A2** shows the age effects obtained from cross-sectional analyes (*β*_CS_ in model A1, highlighted in lightblue) and longitudinal analyses (*β*_L_ in model A2, highlighted in red), where negative coefficients indicate that older age links to decline in trait values. **Panel A3** shows the birth year effects obtained from model A3 (*β*_C_, in cyan), where negative coefficients indicate that later birth year links to lower trait values. **Panel A4** shows the non-linear age effects (*β*_Q_ in model A4, in orange). **Panel A5** contrasts the baseline participation adjusted cross-sectional age effects (*β*_W_, obtained from model A5, in darkblue) to cross-sectional age effects obtained from the UKB baseline sample (*β*_CS_ in model A1, highlighted in lightblue) and the longitudinal UKB sample (in green). The error bars represent the 95% confidence intervals. The bar plot to the left indicates how much of the variance (in %) in effect size differences between longitudinal and cross-sectional age effects (*β*_*L*_− *β*_*CS*_) was explained by cohort effects (in cyan), non-linear age effects (in orange) and selective participation (in darkblue). The source data used in this figure can be found in Supplementary Data 1.

To investigate potential sources of bias underlying these discrepancies, we examined three possible contributors, including birth cohort effects (A3, up to *N* = 99,459), non-linear age effects (A4, up to *N* = 19,374) and selective participation (A5, up to *N*_EFF_= 141,147). Significant Bonferroni-corrected cohort effects (discovery threshold: *P* < 0.05/ 31) were detected for 28 out of the 31 assessed traits (**Figure 2**, model A3). Because date of birth and cross-sectional age are highly inversely correlated in the UKB (*r* = −0.99), negative cohort effects systematically inflate cross-sectional age estimates and, in cases of strong cohort confounding, may even reverse the direction of the observed age effect. For example, substantial negative cohort effects for smoking and alcohol use explained why these traits, which decline with age when assessed longitudinally, falsely appeared to increase with age when tested in cross-sectional analyses. Overall, cohort effects accounted for 90.8% of the variance in discrepancies between longitudinal and cross-sectional estimates (**Figure 2**), thus representing the primary driver of effect-discrepancies between the two designs.

Significant quadratic age effects were detected for 17 traits (*β*_Q_≠ 0 at *P* < 0.05/ 31, **Figure 2**, model A4), indicating that aging trajectories deviated from linearity for some phenotypes. In such cases, models assuming linearity, whether cross-sectional or longitudinal, produce biased estimates, either underestimating or overestimating age effects at the extremes of the age spectrum. Because the age ranges differ between the cross-sectional and longitudinal UKB samples (e.g., a maximum baseline age of 69 years versus 86 at follow-up), the estimated age slopes may differ when non-linear effects are present (c.f., **Supplementary Figure 1** for illustration). However, the relative importance of quadratic age effects appeared to be negligible, accounting for 0.9% (*P* = 0.015) of the variance in effect size differences between designs.

Finally, we assessed the influence of sample representativeness by comparing the crosssectional age effects across three subsamples: a baseline sample adjusted for selective participation (most representative), the unweighted baseline sample, and the longitudinal follow-up sample (least representative). Traits for which the UKB sample was the least representative produced more attenuated cross-sectional age-effect estimates, with sign reversals observed for 4 traits (**Figure 2**, model A5). Although sample representativeness significantly predicted discrepancies between cross-sectional and longitudinal estimates, its contribution was only small compared to the bias induced by the birth year effect (explained variance: 2.8% vs. 90.8%, respectively).

### Cross-sectional versus longitudinal age-varying genetic effects

We identified 57 LD-independent, Bonferroni-corrected variants with significant age-varying effects. The majority of these were detected in cross-sectional analyses (*k*_SNPs_ = 44, up to 406,226 individuals), with fewer reaching significance in longitudinal analyses (*k*_SNPs_ = 14, up to 83,579 individuals). Most of the identified variants were linked to anthropometric traits (weight, body fat–free mass, BMI), metabolic indicators (basal metabolic rate), and clinical outcomes (number of medications used, number of cancers) (**Figure 3B**). In contrast, selfreported health behaviors (e.g., physical activity, smoking, sleep) exhibited no or only few significant age-varying genetic effects. The source data used in **Figure 3B** is provided in **Supplementary Data 2**.

**Figure 3.**
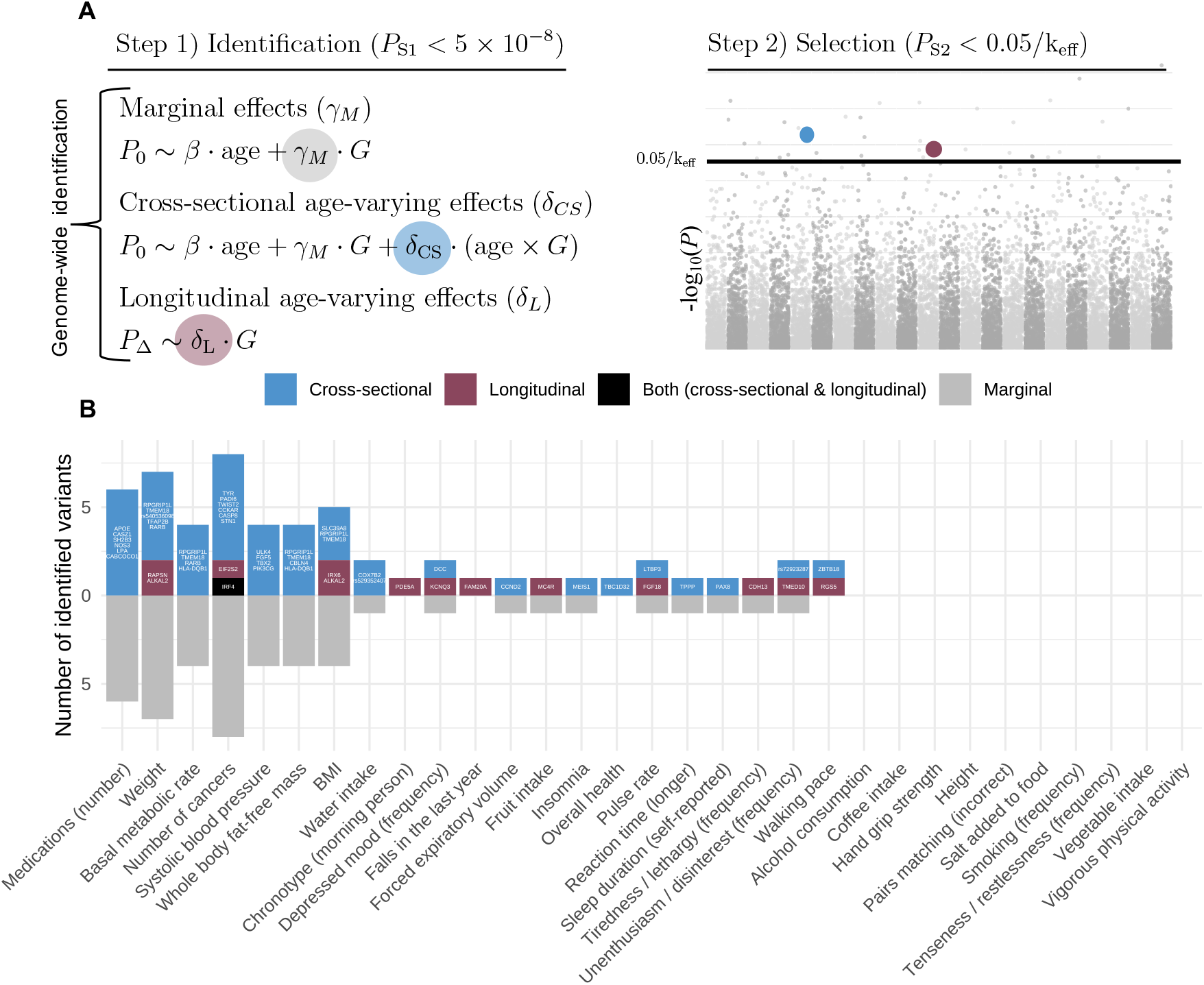
Cross-sectional and longitudinal identification of age-varying genetic effects. **Panel A** describes the two-step genome-wide approach combining marginal (in grey), cross-sectional (in lightblue), and longitudinal (in red) models to detect age-varying genetic effects. In step one, we performed genome-wide scans for models on marginal genetic effects (*γ*_*M*_), crosssectional gene-by-age interaction effects (*δ*_CS_) and genetic effects on change (*δ*_L_). In step two, we selected all LD-independent SNPs reaching genome-wide significance (*P* < 5 × 10^−8^) in either of the three models as candidate genes. We then selected all Bonferroni-corrected significant variants (discovery threshold: *P*_*S*2_< 0.05/*k*_Eff_) from the two models testing for age-varying genetic effects (*δ*_CS_, *δ*_L_), where *k*_Eff_ corresponds to the total number of LD-independent variants reaching genome-wide significance per trait identified in step 1. Annual age-related change (P_Δ_) is derived based on the difference between the standardized phenotype assessed at baseline (*P*_0_) and the most recent follow-up assessment (*P*_1_), divided by the follow-up time. *G* is the genetic variant. The estimated age-varying genetic effects (*δ*) can be interpreted as the genetic effect on annual change. **Panel B** lists all Bonferroni-corrected significant (*P*_S2_< 0.05/k_eff_) genetic variants (mapped to closest gene) identified in either cross-sectional gene-by-age interaction tests (in lightblue), longitudinal tests of age-varying genetic effects (in red) or in both sets of analyses (black). The grey bars at the bottom indicate the number of Bonferroni-corrected significant variants with indicated age-varying genetic effects that were also significant in tests of marginal effects. The source data used in Panel B can be found in Supplementary Data 2.

Regarding the direction of effects, most of the 57 identified variants (85.96%) showed consistent interaction directions across the cross-sectional and longitudinal model. Among those variants, both attenuation and intensification were common modes of interaction (48.98% and 36.73%, respectively) (**Figure 4A**). This pattern appeared to be trait-specific: genetic effects on obesogenic traits tended to attenuate with age, whereas traits adversely impacted by aging (e.g., number of cancers, medication burden, reaction time) more often showed intensifying genetic effects over time. In contrast, cross-over effects were rare (14.29%), with only seven variants exhibiting age-varying effects without detectable marginal effects. In those instances, the direction of the genetic association reversed over time (see panel highlighted in blue, **Figure 4B**, for illustration).

**Figure 4.**
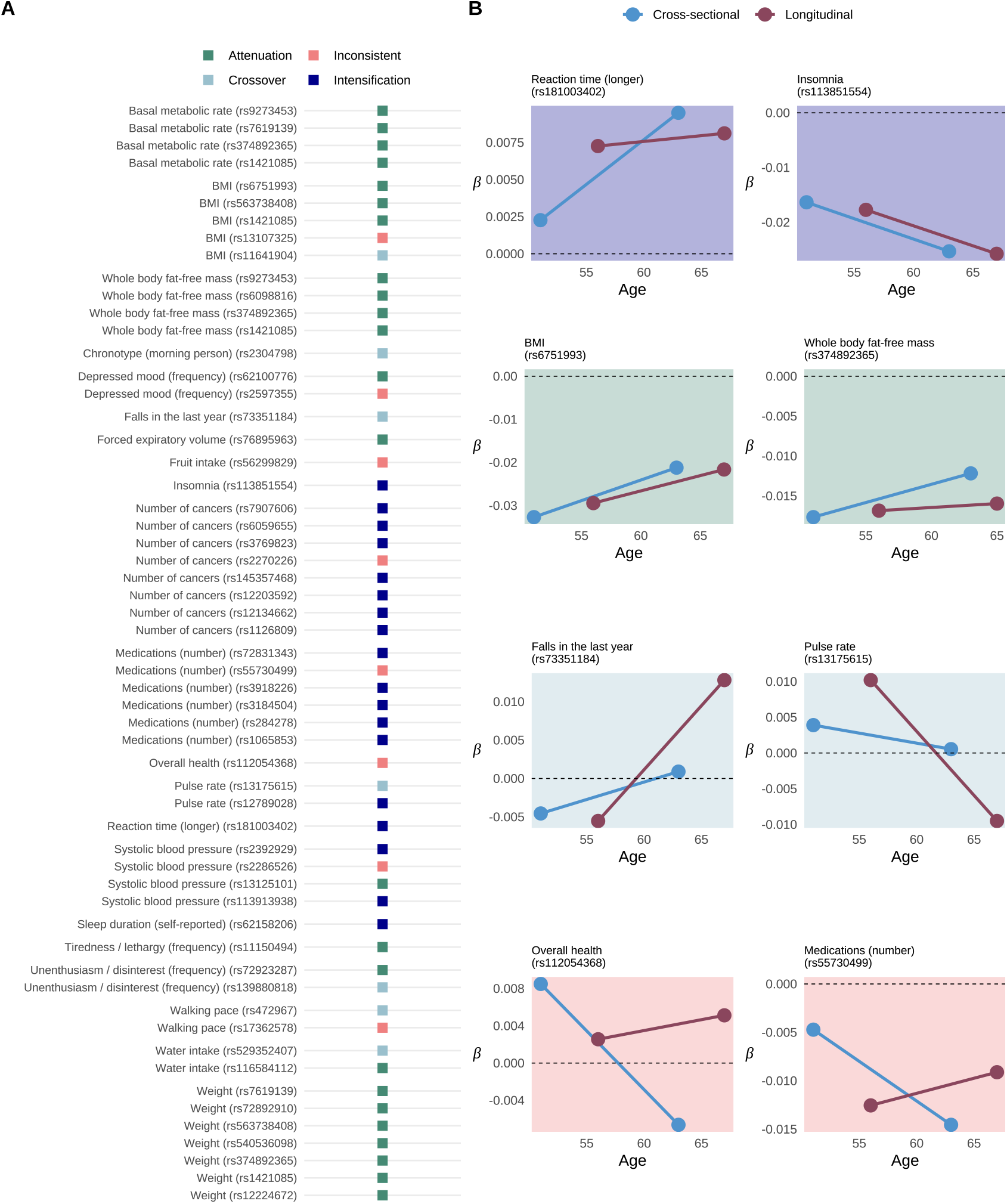
Cross-sectional and longitudinal interpretations of age-varying genetic effects. **Panel A** lists all Bonferroni-corrected significant variants identified in the cross-sectional and/or longitudinal analysis. The colour scheme refers to the interpretation of the age-varying genetic effects, which was inferred based on the direction of the main genetic effect (*γ*_*M*_) and the age-varying genetic effect (*δ*) for a given variant, inferring either attenuation (main and age-varying effect have opposite signs, in lightgreen), intensification (main and age-varying effect have the same direction, in blue) or cross-over (significant age-varying effect are present without a significant marginal effect, in lightblue) of genetic effects over time. Variants highlighted in coral are those with inconsistent direction of age-varying genetic effects (i.e., opposite sign) obtained from the cross-sectional and longitudinal approach. **Panel B** visually illustrates the mechanim of interaction, by plotting the main genetic effect (in standardized coefficients) obtained in younger versus older age groups selected from the baseline UKB sample (splitting individuals around the median age, in lightblue) or the prospective UKB sample (using the baseline and follow-ip assessment, in red). The x-axis indexes the mean age of the sample used to estimate the genetic effects.

While the direction of age-varying effects was largely consistent between cross-sectional and longitudinal designs, the magnitude of effects showed less agreement. Although a significant linear relationship was observed between cross-sectional (*δ*_CS_) and longitudinal (*δ*_L_) estimates (*α*_1_ = 0.71, *P* = 5.60e-11; obtained from *δ*_L_~ *α*_0_ + *α*_1_ ⋅ *δ*_CS_), the slope was significantly different from one (t = −3.35, *P* = 0.0015), indicating that cross-sectional estimates were systematically inflated compared to the longitudinal estimates (see **Supplementary Figure 2**). To better understand these discrepancies, we next examined potential sources of bias, focusing on cohort-varying genetic effects (**Figure 5**, model B3 in up to *N* = 69,178), nonlinear age-varying genetic effects (model B4, up to *N* = 13,851) and selective participation (model B5, in up to *N*_EFF_= 102,148). The source data used in **Figure 5** is provided in **Supplementary Data 3**.

**Figure 5.**
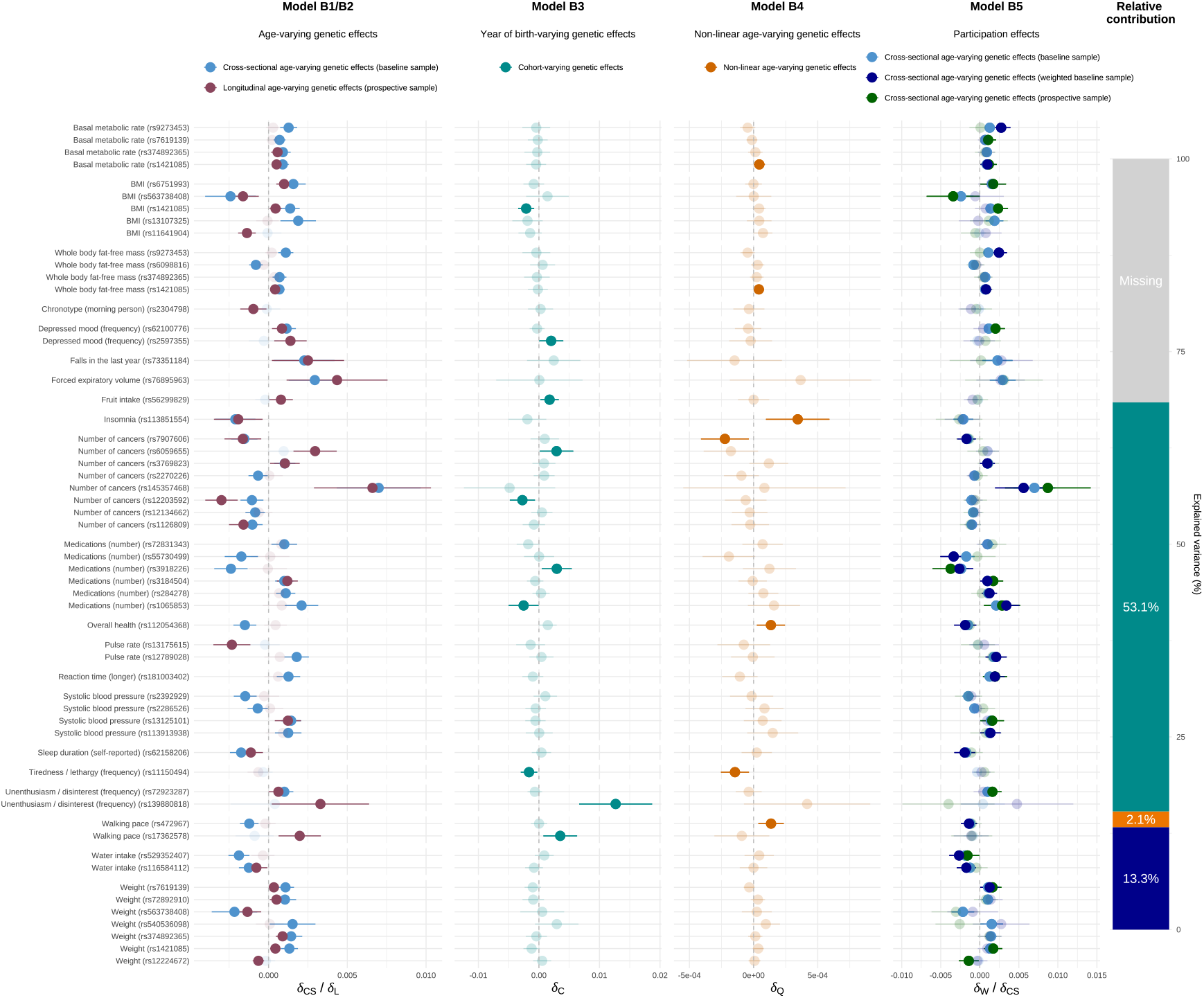
Sources of bias affecting estimates of age-varying genetic effects. **Panel B1/B2** shows the age-varying genetic effects obtained from cross-sectional analyes (*δ*_CS_ in model B1, highlighted in lightblue) and longitudinal analyses (*δ*_L_ in model B2, highlighted in red). **Panel B3** shows the cohort-varying genetic effects obtained from model B3 (*δ*_C_, in cyan). **Panel B4** shows the non-linear (quadratic) age-varying genetic effects (model B4, *δ*_Q_, in orange). **Panel B5** contrasts the baseline participation adjusted cross-sectional age-varying genetic effects (obtained from model B5, *δ*_W_, in darkblue) to crosssectional age-varying genetic effects obtained from standard regression analysis in the UKB baseline sample (in lightblue) and the UKB longitudinal sample (in green). The error bars represent the 95% confidence intervals. Estimates shown in more saturated colors indicate nominally significant effects (*P* < 0.05). The bar plot to the left indicates how much of the variance (in %) in the discrepancies between longitudinal and cross-sectional estimates (*δ*_L_− *δ*_CS_) was accounted for by cohort-varying genetic effects (in cyan), non-linear age-varying genetic effects (in orange) and selective participation (in darkblue). The source data used in this figure can be found in Supplementary Data 3.

Suggestive gene-by-cohort effects (that is, genetic effects that vary across the year of birth) were observed for 10 of the 57 examined variants (*δ*_C_≠ 0 at *P* < 0.05). Together, these year-of-birth-dependent effects explained 53.1% of the variance in discrepancies between cross-sectional and longitudinal estimates, representing the principal source of bias in crosssectional analyses. Notably, for several variants (e.g., rs2597355 on depression, rs56299829 on fruit intake, rs17362578 on walking pace), confounding due to year of birth interactions resulted in a reversal of the estimated direction of age-varying genetic effects when using cross-sectional models.

We did find some evidence of non-linear age-varying genetic effects for 7 of the 57 examined variants (*δ*_Q_≠ 0 at *P* < 0.05). While this phenomenon impacts both cross-sectional and longitudinal interaction estimates, it does not necessarily lead to discrepancies. In line with this argument, deviations from linearity represented only a negligible source of effect-discrepancies between the two designs, accounting for 2.1% of the variance. Of note, because these estimates are derived from smaller subsets of the UK Biobank and are thus subject to greater statistical larger estimation error, the variance component may be underestimated due to regression dilution^35^. To quantify this bias, we calculated the dilution ratio (*λ* ∈ [0, 1]), where lower values denote greater attenuation. For the three factors considered (cohort, age^2^, participation), non-linear age-varying effects exhibited the largest dilution (*λ* = 0.35). After correction for attenuation bias 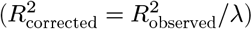, the variance explained by non-linear age effects increased from 2.1% to 5.63% (**Supplementary Figure 3** and corresponding Supplementary Note).

More relevant in explaining effect-discrepancies was bias due to selective participation (13.3% explained variance). Although selection bias can affect both designs, the nature of this bias may differ between approaches. Cross-sectional estimates are particularly affected by selective participation at baseline, which can impact findings if the initial sample is non-representative. In contrast, longitudinal models account for such time-invariant selection mechanisms (e.g., baseline phenotype levels), but assume that continued participation is independent of timevarying factors (e.g., changes in phenotype)^41^. Because different selection pressures may act during the initial recruitment and follow-up, participation bias can contribute to discrepancies between cross-sectional and longitudinal results.

## Discussion

Identifying age-varying genetic effects is essential for advancing our understanding of the dynamic processes characterizing genetic influences on complex traits. Both cross-sectional and longitudinal approaches are designed to tackle this question, but come with distinct strengths and limitations. In this work, we systematically evaluated the degree of convergence between estimates of age-dependent trait changes and age-dependent genetic effects derived from cross-sectional and longitudinal approaches, and investigated key sources of bias that may underlie any observed discrepancies.

At the phenotypic level, cross-sectional age effects deviated from longitudinal estimates, with around 71% of the variance shared between the two approaches. The largest discrepancies were observed for behavioral and lifestyle traits such as smoking and alcohol use, mainly due to confounding by birh year (i.e., cohort) effects, which can substantially alter or even reverse cross-sectional age estimates.

At the genotypic level, we observed high levels of consistency between cross-sectional and longitudinal approaches with respect to the direction of age-varying genetic effects. In both designs, attenuation, characterized by a decrease in genetic effects with advancing age, was a common pattern of interaction particularly for metabolic and well-being traits. This observation aligns with prior studies reporting decreasing heritability with advancing age during adulthood, as documented for BMI^42^ or depression^43^. Such pattern suggest that accumulation of environmental exposures and age-related lifestyle changes (e.g., medication use, changes in diet, sleep, occupational status) likely gain importance for those traits, thereby diminishing the relative contribution of genetics.

Under equivalence, cross-sectional and longitudinal analyses are expected to produce identical estimates of age-varying genetic effects. However, we observed only moderate agreement in the magnitude of these effects, suggesting that between-individual age differences may not accurately reflect within-individual age-related change. Among the sources of bias considered, gene-by-cohort effects stood out as the main contributor, explaining the largest proportion of variance in effect discrepancies. Such effects pose a particular challenge for cross-sectional designs, which are unable to isolate the effects of biological aging from historical or societal changes that differentially impact birth cohorts. Of note, while we have primarily considered cohort effects as a source of bias in the estimation of age-dependent genetic effects, it is important to recognize that such effects also represent meaningful and interpretable variation, capturing how historical or societal changes influence genetic effects across generations. This underscores the value of future research explicitly modeling gene-by-cohort interactions, which may provide important insights into contextual factors that shape genetic risk.

Selective participation was identified as the second most relevant source of bias, leading to differences in age-varying genetic effects obtained from the two designs. Although longitudinal designs are often regarded as more robust due to their ability to adjust for participation mechanisms unrelated to change^41^ (e.g., initial selection into the sample), they are susceptible to selective dropout and survival effects^18,44^. In contrast, UKB cross-sectional analyses are affected by the well-documented healthy volunteer bias present at baseline^32,45–47^. Since the mechanisms driving participation at recruitment may differ from those affecting continued study involvement, these distinct selection pressures can contribute to discrepancies between cross-sectional and longitudinal results. It is important to note, however, that the bias observed here is most relevant to samples requiring active baseline and follow-up participation and may not generalize to prospective studies relying on passive data collection, such as those using routinely collected electronic health records.

Finally, we found that deviations from linearity contributed minimally to the differences in effect estimates between the two designs. Nevertheless, testing for non-linear age effects remains important for two reasons. First, if non-linear trajectories are present but not considered, both cross-sectional and longitudinal models assuming linearity will produce inaccurate estimates. Exploration of possible non-linear patterns can therefore improve model specification and interpretability of the results. Second, when age-related change is non-linear, the estimated linear slope becomes dependent on the underlying age distribution of the sample (see **Supplementary Figure 1**). Because the age ranges differ between the cross-sectional and longitudinal UKB samples (for example, a maximum baseline age of 69 years versus 86 at follow-up), the observed age slopes are not directly comparable across designs, potentially leading to discrepant findings.

Overall, our findings highlight that longitudinal and cross-sectional designs are characterized by distinct strengths and limitations. Longitudinal cohort-heterogeneous designs allow for direct modeling of within-individual change over time, the identification of gene-by-cohort effects, and the detection of non-linear trajectories—insights that cannot be generated by cross-sectional designs but that are crucial for drawing valid conclusions about age-dependent effects. Conversely, cross-sectional analyses provided greater statistical power in our analyses of the UKB data, owing to their larger sample sizes and wider age ranges, thereby facilitating discovery of potentially age-varying genetic effects. Noteworthy, this power advantage is specific to the current dataset, as longitudinal data characterized by minimal attrition and frequent follow-up assessments may offer comparable or even greater statistical power, while preserving the ability to model change over time. Ultimately, neither design is without limitations, and the optimal choice will depend on the specific research question, data availability, and tolerance for different sources of bias. Ideally, researchers may integrate both longitudinal and cross-sectional approaches, which offers a computationally feasible and more robust framework for identifying, quantifying, and mitigating potential threats to the validity of dynamic genetic effects across the lifespan.

### Methodological considerations and analytical constraints

A central assumption of our analytical framework is that period effects−changes attributable to the specific calendar year in which individuals are assessed−are negligible. Such effects can bias age-related estimates if societal changes (e.g., economic crises, public health interventions) affect all age groups simultaneously, and would lead to misattribution of cohort effects in our longitudinal models. Because age, cohort, and period are exactly collinear (age = period − cohort, i.e., the “identification problem”^48^), it is statistically impossible to estimate all three effects simultaneously without constraints, typically by setting period effects to zero^49,50^. Overall, we believe the cohort effects identified in this study represent genuine cohort-specific influences rather than misattributed period effects. Theoretically, cohort effects capture exposures and experiences during formative developmental periods, which are likely to exert more lasting impacts on genetic associations than contemporaneous influences later in life. Supporting this, most gene-by-cohort effects detected here were observed for obesogenic traits and health behaviours, plausibly reflecting generational influences stemming from the 20th-century obesity epidemic, alongside improved public health initiatives and medical care^51–53^. Empirically, we conducted sensitivity analyses excluding participants whose follow-up occurred during or following the COVID-19 pandemic, the most significant recent environmental shock. Results showed high concordance with the full sample for both phenotypic (*r* = 0.98) and genotypic (*r* = 0.98) associations (**Supplementary Figure 3**), suggesting that even substantial period disruptions had minimal impact on our findings.

## Data availability

The genome-wide summary statistics generated as part of this work will be deposited in the GWAS Catalog upon publication of this work.

## Code availability

The following software was used to run the analyses:

- REGENIE^36^ (https://github.com/rgcgithub/regenie)
- *bigsnpr*^54^(https://privefl.github.io/bigsnpr/)
- *gwasrapidd*^55^(https://rmagno.eu/gwasrapidd/)
- *otargen*^56^(https://amirfeizi.github.io/otargen/)
- *survey*^31^(CRAN)
- *lme4*^24^(CRAN)

All analytical scripts are available at https://github.com/TabeaSchoeler/TS2024_ageEffect

## Acknowledgements

This research has been conducted with the UK Biobank Resource under application number 16389; we thank all biobank participants for sharing their data. The computations were performed on the High-Performance Computing Center of University of Lausanne, Switzerland. Z.K. was funded by the Department of Computational Biology of the University of Lausanne and the Swiss National Science Foundation (#310030-189147, #315230-219587). SW and TWW were funded by the Deutsche Forschungsgemeinschaft (DFG, German Research Foundation)—Project-ID 509149993, TRR 374.

