## Supplementary File for "Design and model choices shape inference of age-varying genetic effects on complex traits"

### Supplement File

#### Contents

**Supplementary Figures**

Supplementary Figure 1. Illustration of non-linear age effects . . . . . . . . . . . . . . . . . . . . . . . .

Supplementary Figure 2. Consistency of age-varying genetic effects obtained from cross-sectional and longitudinal analyses . . . . . . . . . . . . . . . . . . . . . . . . . . . . . . . . . . . . . . . . .

Supplementary Figure 3. Quantifycation of regression dilution bias . . . . . . . . . . . . . . . . . . . .

Supplementary Figure 4. Age-varying genetic effects obtained in individuals participating prior to the COVID-19 pandemic . . . . . . . . . . . . . . . . . . . . . . . . . . . . . . . . . . . . . . . . . .

### Supplementary Figures

#### Supplementary Figure 1. Illustration of non-linear age effects

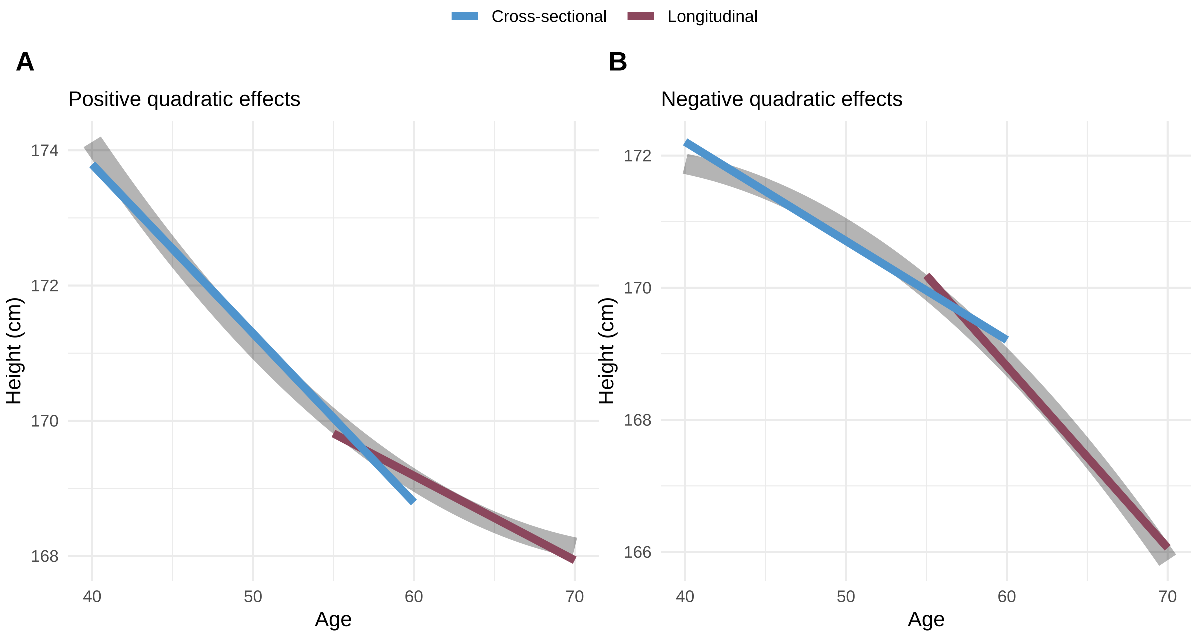


**Supplementary Figure 1** illustrates the impact of non-linear (quadratic) age effects on cross-sectional and longitudinal slope estimates. The figure depicts simulated age trajectories for an outcome variable (e.g., height) as a function of age, including both linear and quadratic (nonlinear) age effects. The underlying outcome is modeled as height $= 170 + \beta_1 \cdot \text{age}_c + \beta_1 \cdot \text{age}_c^2$, where $\beta_1$ is a linear age effect, $\beta_2$ is a quadratic age effect (set to either positive or negative for illustrative purposes) and $\text{age}_c$ denotes age centered at the mean. Two age ranges were selected to represent different cross-sectional (ages 40–60) and longitudinal (ages 55–70) study windows. Linear regression slopes are fit separately to each age window, representing the estimated age effect from cross-sectional (in blue) and longitudinal (in red) approaches, respectively. The plot highlights how the estimated slope (i.e., age effect) depends on the underlying age distribution when non-linear effects are present, leading to potentially discrepant estimates between study designs.

**Supplementary Figure 2. Consistency of age-varying genetic effects obtained from cross-sectional and longitudinal analyses**

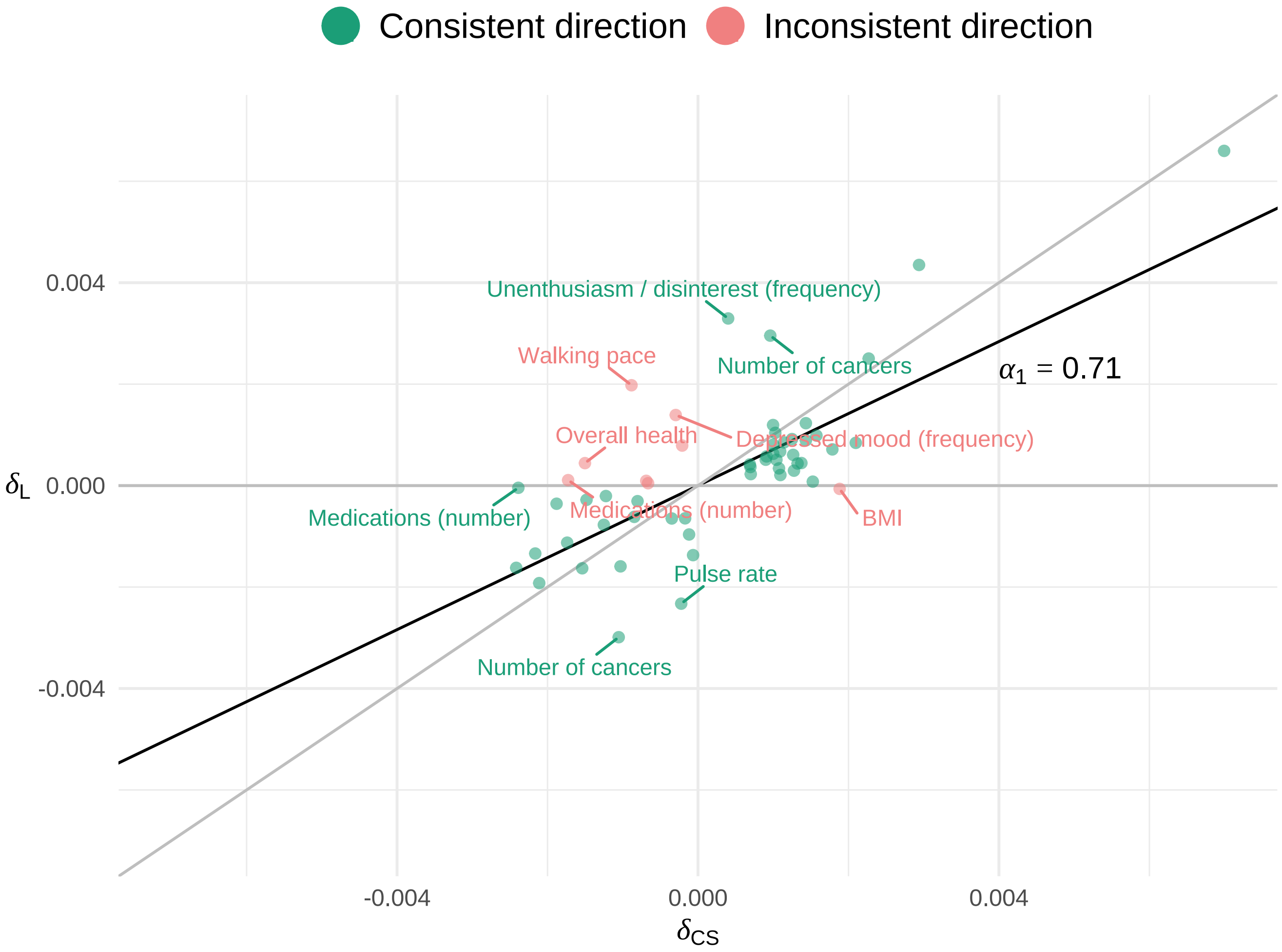


**Supplementary Figure 2** plots the effect sizes of all identified variants with age-varying effects obtained from cross-sectional analyses ($\delta_{\mathrm{CS}}$, on the x-axis) and longitudinal models ($\delta_{\mathrm{L}}$, on the y-axis). The regression coefficient ($\alpha_1$) was obtained from the model $\delta_{\mathrm{L}} \sim \alpha_0 + \alpha_1 \cdot \delta_{\mathrm{CS}}$, were $\alpha_0$ denotes the intercept. The results show that there was a significant linear relationship between the cross-sectional and longitudinal age-varying effects ($\alpha_1 = 0.71$, $P = 5.60\text{e-}11$). However, the slope was significantly different from one (t = -3.35, $P =$ 0.001), indicating that cross-sectional estimates were systematically inflated compared to the longitudinal estimates.

#### Supplementary Figure 3. Quantifycation of regression dilution bias

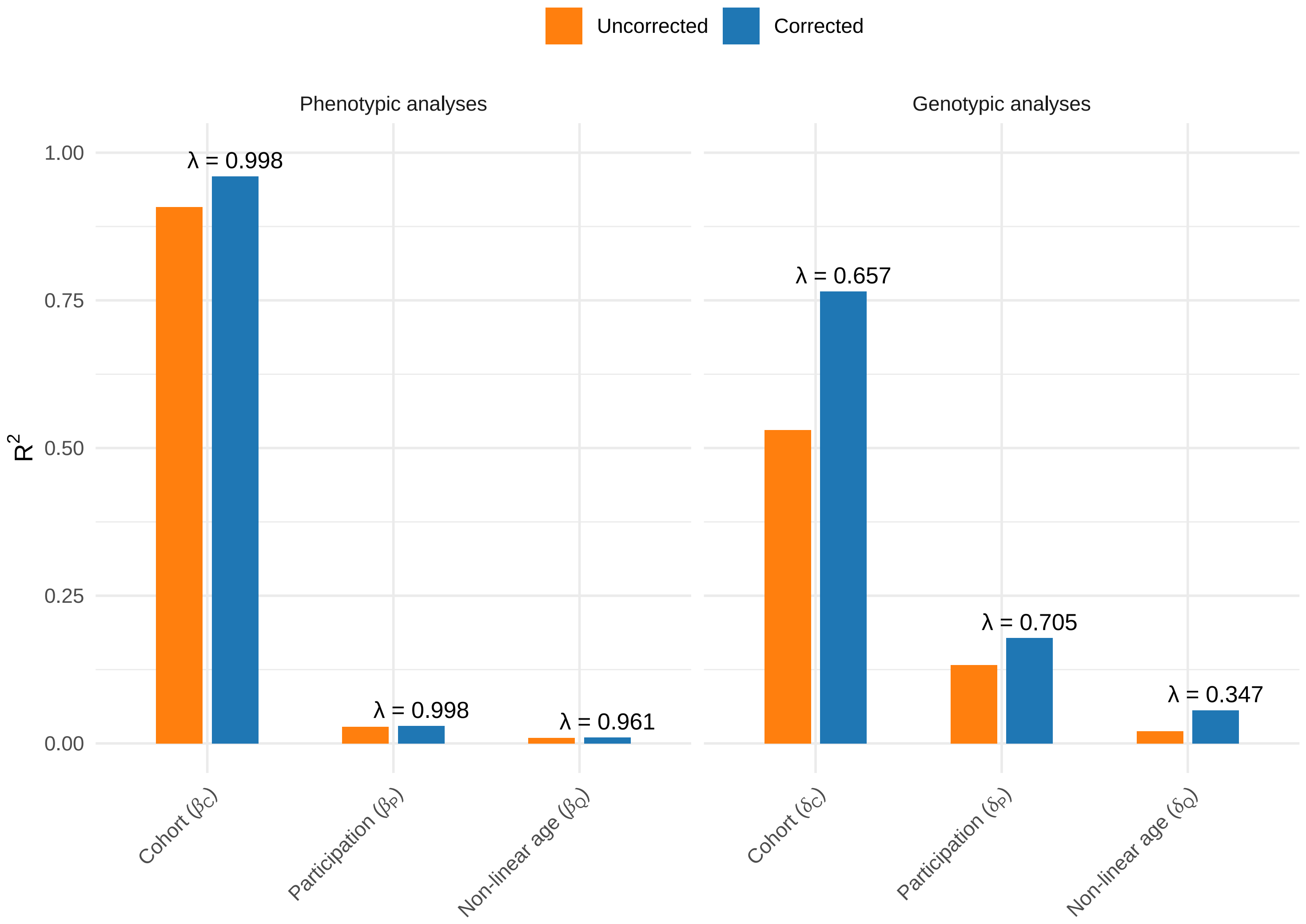


The figure shows the uncorrected (in in orange) and attenuation bias corrected (in in blue) $R^2$-contribution per factor (cohort effects, non-linear age effects, participation bias) to the observed effect differences between the cross-sectional and longitudinal design. As described and plotted in **Figure 2** and **Figure 5** in the main manuscript, the uncorrected $R^2$-contributions were obtained from two sets of analyses:

(1) Phenotypic analyses (**Figure 2**): $\beta_{\mathrm{L}} - \beta_{\mathrm{CS}} \sim \alpha_0 + \alpha_1 \cdot \beta_{\mathrm{C}} + \alpha_2 \cdot \beta_{\mathrm{Q}} + \alpha_3 \cdot \beta_{\mathrm{P}}$

(2) Genotypic analyses (**Figure 5**): $\delta_{\mathrm{L}} - \delta_{\mathrm{CS}} \sim \alpha_0 + \alpha_1 \cdot \delta_{\mathrm{C}} + \alpha_2 \cdot \delta_{\mathrm{Q}} + \alpha_3 \cdot \delta_{\mathrm{P}}$

We corrected for attenuation bias by dividing the observed variance components by a correction factor $\lambda$, such that $R^2_{\mathrm{corrected}} = R^2_{\mathrm{observed}}/\lambda$. Here, $\lambda$ is the dilution ratio ($\lambda \in [0, 1]$), which quantifies the proportion of total observed variance in regression coefficients that reflects true underlying effects: $\lambda = \frac{V_{\mathrm{total}} - V_{\mathrm{error}}}{V_{\mathrm{total}}}$, with

$V_{\text{total}} = \frac{1}{n-1}\sum_{i=1}^{n}(\hat{\theta}_i - \mu)^2$ and $V_{\text{error}} = \frac{1}{n}\sum_{i=1}^{n} SE(\hat{\theta}_i)^2$. $\hat{\theta}$ denotes the vector of estimated coefficients for a given factor (e.g., $\hat{\theta}_i = \hat{\beta}_{\text{C}_i}$ for cohort effects) and $\mu = \frac{1}{n}\sum_{i=1}^{n}\hat{\theta}_i$. Lower values of $\lambda$ indicate greater measurement error and stronger attenuation bias. The corrected $R^2$-estimates were rescaled to sum up to 1.

Of note, participation effects were defined as the difference between estimates obtained from a representative (weighted, W) and a less representative (unweighted, U) sample ($\hat{\theta}_{\text{P}} = \hat{\theta}_{\text{W}} - \hat{\theta}_{\text{U}}$). The corresponding standard errors were obtained as: $SE(\hat{\theta}_{\text{P}}) = \sqrt{SE(\hat{\theta}_{\text{W}})^2 + SE(\hat{\theta}_{\text{U}})^2 - 2r \cdot SE(\hat{\theta}_{\text{W}}) \cdot SE(\hat{\theta}_{\text{U}})}$, where $\hat{\theta} \in \hat{\beta}, \hat{\delta}$ represents either phenotypic or genotypic estimates. To obtain $r$, we performed simulations using data from 80,000 UK Biobank participants with available sampling weights and taking part in follow-up assessments. In 1,000 iterations, we generated normally distributed variables $X$ and $Y$, and fit two linear models per iteration: (1) a weighted regression using normalized sampling weights, and (2) an unweighted regression with all weights set to 1. The correlation between the regression coefficients obtained from $Y \sim X$ across iterations was used to approximate $r$. This yielded an estimated value of $r = 0.6$.

**Supplementary Figure 4. Age-varying genetic effects obtained in individuals participating prior to the COVID-19 pandemic**

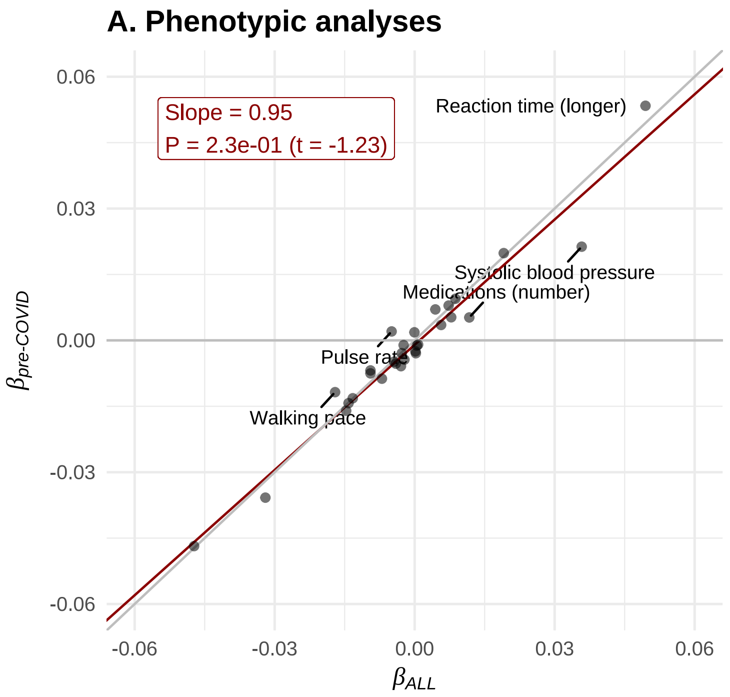


**Supplementary Figure 3A** plots the longitudinal age effects ($\beta$) estimated for all 31 included traits (obtained from $P_{tj} = \alpha_0 + \beta \cdot \text{age}_{c,tj} + u_j + \varepsilon_{tj}$, see model A1 in the main manuscript). Estimates are shown for the full UK Biobank sample ($\beta_{\text{ALL}}$) and for a restricted subsample that excludes participants whose follow-up occurred during or after the COVID-19 pandemic (after 31st December 2019; $\beta_{\text{pre-COVID}}$). The text box in the plot shows the slope ($\alpha_1$) from the regression $\beta_{\text{ALL}} \sim \alpha_0 + \alpha_1 \cdot \beta_{\text{pre-COVID}}$, where $\alpha_0$ is the intercept. The associated $P$-value was obtained from a two-sided $t$-test, assessing whether the slope significantly differed from one.

**Supplementary Figure 3B** displays the effect sizes for all variants showing age-varying effects, estimated from longitudinal genome-wide models ($P_{\Delta_j} = \mathbf{W} \cdot \alpha + \delta \cdot G_j + \varepsilon_j$, see main manuscript) using REGENIE. As above, estimates are shown for both the full sample ($\delta_{\text{ALL}}$) and the pre-pandemic subsample ($\delta_{\text{pre-COVID}}$). The text box includes the slope ($\alpha_1$) from the regression $\delta_{\text{ALL}} \sim \alpha_0 + \alpha_1 \cdot \delta_{\text{pre-COVID}}$, and the $P$-value testing if the slope significantly differed from one.

Overall, effect estimates derived from participants assessed before the COVID-19 pandemic were highly concordant with those from the full sample, for both phenotypic ($r = 0.98$, $P = 4.8\text{e-}20$) and genotypic ($r = 0.98$, $P = 2.9\text{e-}37$) associations.
